# Catch-up Routine Immunization to Restore Childhood Immunization Coverage following COVID-induced Declines in Zamboanga Peninsula, Philippines

**DOI:** 10.1101/2022.11.19.22282451

**Authors:** Mary Germeyn D. Punzalan

## Abstract

Due to numerous waves of COVID-19 surge and a more focused government initiative to jump-start the COVID-19 vaccination in 2021, routine immunization was neglected in Zamboanga Peninsula, Philippines, ending the year with a fully-immunized coverage of only 59%. To address this, an intensified catch-up immunization campaign was conducted in April-June 2022 among children 0-23 months of age who missed their routine immunizations. A program case study is utilized to describe the catch-up immunization campaign conducted. The target for the catch-up immunization campaign was to vaccinate at least 80% of infants ages 0-23 months who missed any of their routine immunizations schedule. Online microplanning workshops and consultative meetings were conducted in preparation for the campaign. A record review was conducted from data from municipal and provincial health offices and then consolidated at the regional level. The coverage for all antigens at the regional level (all cities, and municipalities combined), was 49-60%, except for inactivated polio vaccine 2 (32%). In other words, approximately half of those children under 23 months who had missed doses got vaccinated during the catch-up vaccination activities. This was a relatively fair turnout of the campaign considering that the local government units are still transitioning to non-COVID health services. To address the immunization gap, the catch-up immunization campaign will continue to be conducted wherein local government unit health workers have already allotted one day every month to conduct house-to-house vaccination activity to track and vaccinate missed children.

## Introduction

Zamboanga Peninsula, with a total population of 3,834,801, is located in the western part of Mindanao island, Philippines. It has 3 provinces consisting of 67 municipalities, 4 component cities, and 1 highly-urbanized city. It has 1,904 barangays wherein 353 of which belong to geographically isolated and disadvantaged areas (GIDA).

Since the first case of COVID-19 in the Philippines on March 2020, the WHO and UNICEF have already pre-empted the Department of Health on the possible decline of routine immunization, predicting that 2 million Filipino children may miss out on the vaccination due to the ongoing restrictions brought about by the pandemic. In 2019, the fully immunized child (1 dose of BCG, 3 doses of pentavalent vaccine, 3 doses of OPV, and 2 doses of MMR on or before 1 year of age) coverage of the Zamboanga Peninsula Region achieved one of its lowest coverage at 51% after the Dengvaxia controversy which occurred in 2017 wherein an alleged increase risk for dengue severity was reported among children who received the vaccine during the school-based immunization^1^. This eventually led to a measles outbreak in 2019. With intensified campaigns, the FIC coverage increased to 63% in 2020 but once again took a dip at 52% of the target population in the first semester of 2021 which eventually increased to 59% by the end of 2021. This was due to numerous waves of COVID-19 surge and at the same time a more focused government initiative to jump-start the COVID-19 vaccination.

With the threat of a measles resurgence and a polio outbreak, the Department of Health instituted the Measles-Rubella (MR) and Oral Polio Vaccine (OPV) Supplemental Immunization Activity (SIA) from October to November 2020 to interrupt disease transmission^1^. This campaign achieved the desired population immunity of 95.8% among children ages 9-59 months. However, given the need to maintain the desired population immunity to prevent outbreaks, catch-up routine immunization activities began on October 2021.

However, despite streamlining the guidelines for routine catch-up immunization, COVID-19 vaccination and routine immunization efforts shared and competed for the same resources (e.g., human resources, logistics, providers, commodities, and local government unit commitment, leading to lower routine immunization coverage. When the Zamboanga Peninsula eventually reached almost 70% of COVID-19 fully vaccinated in April 2022, the public’s non-COVID-related health services, including routine immunization, were given recognition and importance by the public. Utilizing the success of the National COVID-19 Vaccination Days wherein multi-agency policies were set forth to address supply needs and gaps in demand generation, a similar intensity of the governmental and societal actions were incorporated into the catch-up of under-immunized vulnerable children. Called “*Chikiting Bakunation Days: National Vaccination Days for Catch-up and Routine Immunization*”, approaches taken during the National COVID-19 Vaccination Days were adopted for catch-up immunization^2^. This report describes the catch-up immunization activities in the Zamboanga Peninsula in April-June 2022 among children 0-23 months of age who missed their routine immunizations.

### Program Description

On October 2021, an online consultative meeting with the National Immunization Program (NIP) Manager from the Department of Health-Disease Prevention and Control Bureau (DOH-DPCB) was conducted with representative NIP coordinators at the sub-national/regional DOH offices to cascade the guidelines for the implementation of the routine catch-up immunization.

At the regional level, the guidelines were cascaded to the NIP coordinators from the 3 provinces (Zamboanga del Sur, Zamboanga del Norte, and Zamboanga Sibugay) and 5 cities (Zamboanga City, Pagadian City, Isabela City, Dipolog City, and Dapitan City). Because of COVID movement restrictions, microplanning workshops were conducted online. The following activities were conducted at the provincial, city, and municipal levels^1,2^:

1. Review of immunization coverage down to the municipal/city level, including analysis of immunization reports at the barangay level to identify barangays and puroks with a high number of unimmunized children.
2. Identification of children under 23 months of age who missed their primary immunization series through review of master listings, target client lists, and immunization cards.
3. Routine catch-up immunization every last week of the month utilizing the following strategies:
  a. Door-to-door vaccination
  b. Modified fixed posts (e.g., school/barangay gymnasiums, temporary outreach sites in puroks)
  c. Fixed sites (e.g., barangay health station, rural health units, district health centers)
4. Recording and reporting of routine catch-up accomplishment reports and updating the target client list for children who were vaccinated during the activity
5. Vaccine supply chain and management procedures and the surveillance and reporting of adverse events following immunization (AEFI)

Implementation of catch-up immunization on the last week of the month began in November 2021 and was endorsed by the Regional Inter-Agency Task Force on COVID-19 (RIATF), a collaborative partnership and policy-making body of regional government agencies and stakeholders in the Zamboanga Peninsula. The endorsement from the RIATF enabled local chief executives to ensure the implementation of the activity in their areas of responsibility.

Catch-up routine immunization activities were only conducted for a month and were halted when increased Omicron COVID cases resulted in an intensified COVID vaccination from December 2021 through February 2022. All healthcare workers and local government unit resources were focused on the COVID-19 response. When COVID cases decreased in March, the DOH-DPCB proposed to conduct the National Vaccination Days for Routine and Catch-up Immunization from April to June 2022. The last Thursday and Friday of those months were allotted for intensified catch-up immunization activities. The National Vaccination Days for Routine and Catch-up Immunization adopted the successful governmental and societal approach of the National Vaccination Days for COVID-19, and the approach was adopted at the regional level. Two (2) days were allotted every month from April to June 2022 for intensified catch-up immunization activities.

**Figure 1.**
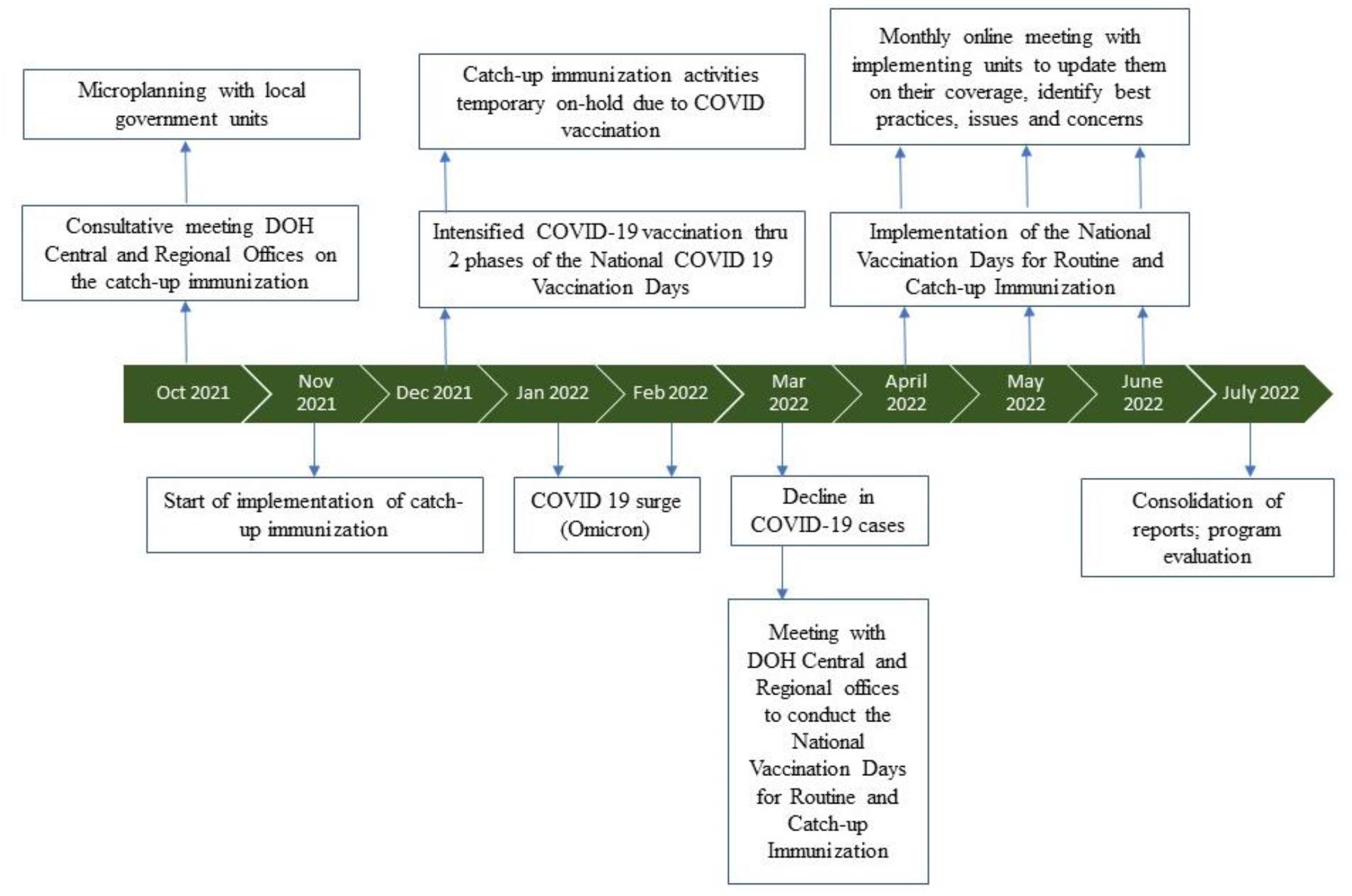
Timeline of activities during implementation of the National Vaccination Days for Catch-up Immunization and COVID surge and COVID vaccination day.

## Methods

A program case study was utilized to describe the catch-up immunization campaign conducted in Zamboanga Peninsula, Philippines. The target for the catch-up immunization campaign was to vaccinate at least 80% of infants ages 0-23 months who missed any of their routine immunizations schedule. The 80% target was set by the Department of Health-Disease Prevention and Control Bureau (DOH-DPCB) and calculated as:

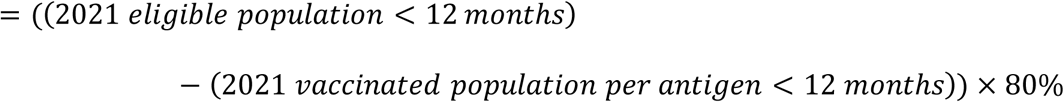

A record review was conducted from data from municipal and provincial health offices and then consolidated at the regional level. Due to the nature of the study, this was granted an exemption for ethics review by the Ateneo de Zamboanga University Research Ethics Committee. During the vaccination activities at the implementing sites, verbal consents were taken from the primary caregivers prior to vaccination.

## Results

Table 1 presents the target number of children with missed doses, the number of doses given during catch-up, and immunization coverage among children with missed doses, per antigen. With a budget of Php 4,382,491 to shoulder mobilization of vaccination teams, vaccine freight, and handling, and commodities (e.g. AD syringes, cotton balls), the coverage for all antigens at the regional level (all cities, and municipalities combined), was 49-60%, except for IPV2 (32%). In other words, approximately half of those children under 23 months who had missed doses got vaccinated during the catch-up vaccination activities.

**Table 1.**
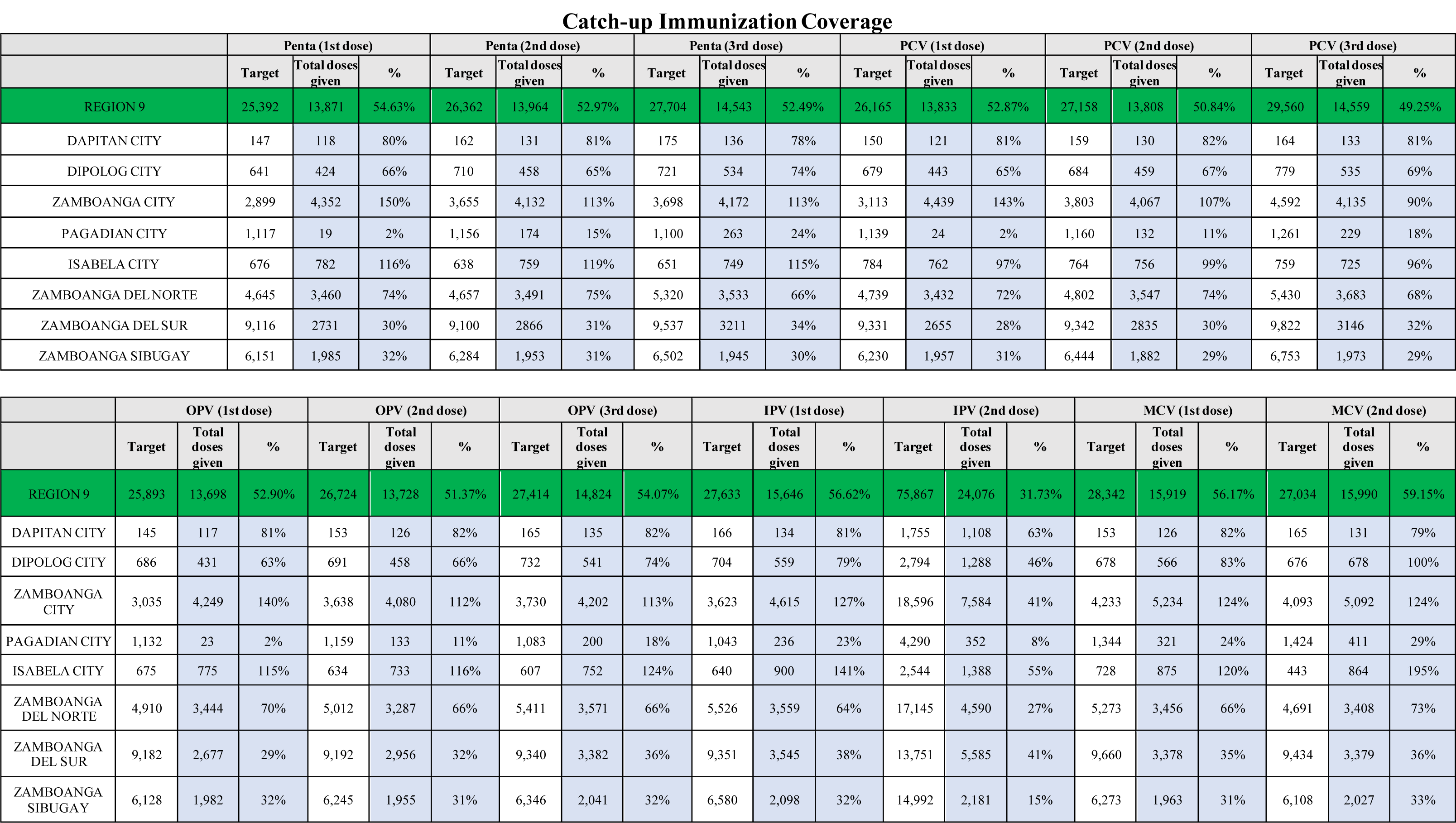
Catch-up immunization coverage among infants ages 0-23 months who missed their routine immunizations by antigen and municipality, Zamboanga Peninsula, April - June 2022.

Catch-up Immunization Coverage
|  | Penta (1st dose) |  |  | Penta (2nd dose) |  |  | Penta (3rd dose) |  |  | PCV (1st dose) |  |  | PCV (2nd dose) |  |  | PCV (3rd dose) |  |  |
| --- | --- | --- | --- | --- | --- | --- | --- | --- | --- | --- | --- | --- | --- | --- | --- | --- | --- | --- |
|  | Target | Total doses given | % | Target | Total doses given | % | Target | Total doses given | % | Target | Total doses given | % | Target | Total doses given | % | Target | Total doses given | % |
| REGION 9 | 25,392 | 13,871 | 54.63% | 26,362 | 13,964 | 52.97% | 27,704 | 14,543 | 52.49% | 26,165 | 13,833 | 52.87% | 27,158 | 13,808 | 50.84% | 29,560 | 14,559 | 49.25% |
| DAPITAN CITY | 147 | 118 | 80% | 162 | 131 | 81% | 175 | 136 | 78% | 150 | 121 | 81% | 159 | 130 | 82% | 164 | 133 | 81% |
| DIPOLOG CITY | 641 | 424 | 66% | 710 | 458 | 65% | 721 | 534 | 74% | 679 | 443 | 65% | 684 | 459 | 67% | 779 | 535 | 69% |
| ZAMBOANGA CITY | 2,899 | 4,352 | 150% | 3,655 | 4,132 | 113% | 3,698 | 4,172 | 113% | 3,113 | 4,439 | 143% | 3,803 | 4,067 | 107% | 4,592 | 4,135 | 90% |
| PAGADIAN CITY | 1,117 | 19 | 2% | 1,156 | 174 | 15% | 1,100 | 263 | 24% | 1,139 | 24 | 2% | 1,160 | 132 | 11% | 1,261 | 229 | 18% |
| ISABELA CITY | 676 | 782 | 116% | 638 | 759 | 119% | 651 | 749 | 115% | 784 | 762 | 97% | 764 | 756 | 99% | 759 | 725 | 96% |
| ZAMBOANGA DEL NORTE | 4,645 | 3,460 | 74% | 4,657 | 3,491 | 75% | 5,320 | 3,533 | 66% | 4,739 | 3,432 | 72% | 4,802 | 3,547 | 74% | 5,430 | 3,683 | 68% |
| ZAMBOANGA DEL SUR | 9,116 | 2731 | 30% | 9,100 | 2866 | 31% | 9,537 | 3211 | 34% | 9,331 | 2655 | 28% | 9,342 | 2835 | 30% | 9,822 | 3146 | 32% |
| ZAMBOANGA SIBUGAY | 6,151 | 1,985 | 32% | 6,284 | 1,953 | 31% | 6,502 | 1,945 | 30% | 6,230 | 1,957 | 31% | 6,444 | 1,882 | 29% | 6,753 | 1,973 | 29% |

|  | OPV (1st dose) |  |  | OPV (2nd dose) |  |  | OPV (3rd dose) |  |  | IPV (1st dose) |  |  | IPV (2nd dose) |  |  | MCV (1st dose) |  |  | MCV (2nd dose) |  |  |
| --- | --- | --- | --- | --- | --- | --- | --- | --- | --- | --- | --- | --- | --- | --- | --- | --- | --- | --- | --- | --- | --- |
|  | Target | Total doses given | % | Target | Total doses given | % | Target | Total doses given | % | Target | Total doses given | % | Target | Total doses given | % | Target | Total doses given | % | Target | Total doses given | % |
| REGION 9 | 25,893 | 13,698 | 52.90% | 26,724 | 13,728 | 51.37% | 27,414 | 14,824 | 54.07% | 27,633 | 15,646 | 56.62% | 75,867 | 24,076 | 31.73% | 28,342 | 15,919 | 56.17% | 27,034 | 15,990 | 59.15% |
| DAPITAN CITY | 145 | 117 | 81% | 153 | 126 | 82% | 165 | 135 | 82% | 166 | 134 | 81% | 1,755 | 1,108 | 63% | 153 | 126 | 82% | 165 | 131 | 79% |
| DIPOLOG CITY | 686 | 431 | 63% | 691 | 458 | 66% | 732 | 541 | 74% | 704 | 559 | 79% | 2,794 | 1,288 | 46% | 678 | 566 | 83% | 676 | 678 | 100% |
| ZAMBOANGA CITY | 3,035 | 4,249 | 140% | 3,638 | 4,080 | 112% | 3,730 | 4,202 | 113% | 3,623 | 4,615 | 127% | 18,596 | 7,584 | 41% | 4,233 | 5,234 | 124% | 4,093 | 5,092 | 124% |
| PAGADIAN CITY | 1,132 | 23 | 2% | 1,159 | 133 | 11% | 1,083 | 200 | 18% | 1,043 | 236 | 23% | 4,290 | 352 | 8% | 1,344 | 321 | 24% | 1,424 | 411 | 29% |
| ISABELA CITY | 675 | 775 | 115% | 634 | 733 | 116% | 607 | 752 | 124% | 640 | 900 | 141% | 2,544 | 1,388 | 55% | 728 | 875 | 120% | 443 | 864 | 195% |
| ZAMBOANGA DEL NORTE | 4,910 | 3,444 | 70% | 5,012 | 3,287 | 66% | 5,411 | 3,571 | 66% | 5,526 | 3,559 | 64% | 17,145 | 4,590 | 27% | 5,273 | 3,456 | 66% | 4,691 | 3,408 | 73% |
| ZAMBOANGA DEL SUR | 9,182 | 2,677 | 29% | 9,192 | 2,956 | 32% | 9,340 | 3,382 | 36% | 9,351 | 3,545 | 38% | 13,751 | 5,585 | 41% | 9,660 | 3,378 | 35% | 9,434 | 3,379 | 36% |
| ZAMBOANGA SIBUGAY | 6,128 | 1,982 | 32% | 6,245 | 1,955 | 31% | 6,346 | 2,041 | 32% | 6,580 | 2,098 | 32% | 14,992 | 2,181 | 15% | 6,273 | 1,963 | 31% | 6,108 | 2,027 | 33% |

The report also showed variations in the coverage rates per province and city. For example, OPV1 coverage was 2% for Pagadian City but 150% for Zamboanga City. MCV2 was 29% in Pagadian City, but 195% for Isabela City. Approximately 50% of the missed children were vaccinated with OPV 1, OPV 2, and OPV 3 but only 31% of the target population received IPV 2. Whereas 56% of missed children received MCV 1 and 59% received their MCV 2 vaccine during the catch-up campaign.

## Discussion

This report reflects the aftermath of the COVID-19 pandemic on routine immunization services in the Zamboanga Peninsula as health workers are slowly shifting their focus to non-COVID services. Local government units (LGUs) reallocated their resources to address the routine immunization gap in their locality. Multi-sectoral collaboration was also evident during the campaign as seen in the support of the Department of Education and Department of Social Welfare and Development in advocating for the immunization activity. The Philippine National Police and the Armed Forces of the Philippines also provided security to vaccination teams going to geographically isolated and disadvantaged areas (GIDA) in the region.

A high accomplishment was noted for Zamboanga City since, being a highly urbanized city, families from the provinces migrate to this city in the hope of greener pastures. Isabela City, also noted to have a high accomplishment, is under a different region called the Bangsamoro Autonomous Region of Muslim Mindanao (BARMM) but when it comes to health and education, Isabela City is under the jurisdiction of Zamboanga Peninsula. Families within the island migrate to Isabela City due to business, trade, and even health care. On the other hand, Pagadian City showed a low accomplishment because the identified population for this city was coming from GIDA barangays known to have very high vaccine hesitancy. The city health office also expressed that their targets were too high and not coinciding with the master list they have.

Considering the history of a polio outbreak in the Zamboanga Peninsula in 2020, it was noted that during this catch-up immunization campaign, approximately 50% of the missed children were vaccinated with OPV 1, OPV 2, and OPV 3. Only 31% of the target population received IPV 2 since, in the Philippines, the second dose administration of IPV started only in 2021 and to date, the allocated IPV doses in the region were not sufficient for the entire <23 months eligible population.

Measles resurgence remains a threat in the Zamboanga Peninsula considering that only 56% of missed children received MCV 1 and 59% received their MCV 2 vaccine during the catch-up campaign. Consequently, this still places the region at risk for future epidemics hence the need to address the immunization gap by intensifying school-based measles vaccination for 6 to 7 years old.

While the direction and implementation of this catch-up immunization campaign stemmed from the success of the COVID-19 vaccination wherein targets were achieved through inter-sectoral collaboration to address supply needs and gaps in demand generation, only half of the set targets were attained due to a highly mobile and transient population with Zamboanga Peninsula and even the BaSulTa (Basilan, Sulu, Tawi-tawi, BARMM) areas. A very high population was also set according to population projections and estimates which did not coincide with the master listing done at the barangay level for most municipalities. The low vaccine confidence^3^ brought about by the Dengvaxia controversy remained an issue that despite intensified social mobilization and demand generation, many still fear the effects of routine immunization. One lesson learned in this campaign was the need for a local chief executive such as a mayor or a governor to be a vaccination champion and enforce policies such as a sustainable form of incentivizing parents and caregivers to bring their children for vaccination.

To address the immunization gap, the catch-up immunization campaign will continue to be conducted wherein local government unit health workers are allotted one (1) day every month to conduct house-to-house vaccination activity to track and vaccinate missed children.

## Key Implications

- The local government commitment through the local chief executives as well as a multi-sectoral approach are important to ensure the success and sustainability of the catch-up immunization campaign and protect vulnerable children from vaccine-preventable diseases.
- To address the immunization gap, the catch-up immunization campaign should be conducted for at least one (1) day every month to conduct house-to-house vaccination activity to track and vaccinate missed children.
- Local chief executives such as a mayors and governors should be lobbied as vaccination champions and enforce policies such as a sustainable form of incentivizing parents and caregivers to bring their children for vaccination.

## Data Availability

The authors declare that the data supporting the findings of this study are available within the article.

